# Clonal Hematopoiesis of Indeterminate Potential and Risk of Major Age-Related Eye Diseases

**DOI:** 10.64898/2026.04.13.26350756

**Authors:** Ruijie Xie, Ben Schöttker

## Abstract

**Importance:** Age-related eye diseases, such as cataract, glaucoma, age-related macular degeneration (AMD), and diabetic retinopathy (DR), are leading causes of irreversible vision loss globally. Chronic inflammation is a shared pathogenic pathway, but the role of systemic inflammatory drivers like clonal hematopoiesis of indeterminate potential (CHIP) is unknown.

**Objective:** To investigate the association of CHIP, including its major genetic subtypes and clone sizes, with the risk of four major age-related eye diseases.

**Design, Setting, and Participants:** This was a prospective cohort study conducted using data from the UK Biobank, a large-scale, population-based cohort. A total of 436,469 participants free of the four eye diseases at baseline were included in the analysis. Data were collected from 2006 to 2010, with follow-up extending to March 2022.

**Exposures:** CHIP status was ascertained from whole-exome sequencing data, defined by the presence of a somatic driver mutation with a variant allele fraction of 2% or greater.

**Main Outcomes and Measures:** The primary outcomes were incident cases of cataract, glaucoma, AMD, and DR, identified through linked electronic health records. Associations were assessed using multivariable Cox proportional hazards regression models.

**Results:** Of 436,469 participants (mean [SD] age, 56.4 [8.1] years; 54.5% women), 14,110 (3.2%) had CHIP. Over a median follow-up of 13.1 years, CHIP was significantly associated with an increased risk of incident cataract (Hazard Ratio [HR], 1.08; 95% CI, 1.03-1.14), AMD (HR, 1.12; 95% CI, 1.04-1.21), and DR (HR, 1.41; 95% CI, 1.20-1.64). No significant association was found with glaucoma (HR, 1.08; 95% CI, 0.99-1.17). The risk for AMD was primarily associated with smaller clones (VAF <10%), while the risk for DR was highest with non-DNMT3A mutations. Systemic inflammation, particularly neutrophil count, partially mediated the associations.

**Conclusions and Relevance:** In this study, CHIP was independently associated with a higher risk of developing cataract, AMD, and DR, but not glaucoma. These findings establish a link between hematopoietic somatic mutations and the pathogenesis of several major age-related eye diseases, suggesting that CHIP-driven inflammation is a potential target for risk stratification and prevention.

**Key Points:** *Question:* Is clonal hematopoiesis of indeterminate potential (CHIP) associated with the risk of major age-related eye diseases?

*Findings:* In this cohort study of 436,469 participants, CHIP was associated with an increased risk of incident cataract (HR, 1.08; 95% CI, 1.03-1.14), age-related macular degeneration (HR, 1.12; 95% CI, 1.04-1.21), and diabetic retinopathy (HR, 1.41; 95% CI, 1.20-1.64), but not glaucoma.

*Meaning:* These findings identify CHIP as an independent, non-ocular risk factor for cataract, AMD, and diabetic retinopathy, suggesting that systemic inflammation driven by CHIP contributes to the pathogenesis of these conditions and may represent a novel target for preventive strategies.

## Introduction

Age-related eye diseases, including cataract, glaucoma, age-related macular degeneration (AMD), and diabetic retinopathy (DR), represent a growing global health crisis. As the leading causes of vision impairment and blindness worldwide, these conditions affect an estimated 2.2 billion people, with much of the associated vision loss being irreversible.^1, 2^ The prevalence of these disorders is projected to increase dramatically with the aging of the global population, posing significant challenges to healthcare systems and quality of life.^3^ Although clinically distinct, these conditions appear to share common pathogenic pathways rooted in chronic, low-grade inflammation and oxidative stress, suggesting that systemic factors capable of modulating inflammation could represent novel, shared risk factors.^4–6^

Clonal hematopoiesis of indeterminate potential (CHIP) is a common, age-related condition characterized by the clonal expansion of hematopoietic stem cells harboring somatic mutations, most often in DNMT3A, TET2, and ASXL1, in the absence of overt hematologic malignancy.^7, 8^ The prevalence of CHIP is strongly age-dependent, rising from less than 1% in individuals younger than 50 years to 10% to 20% in those older than 65 years.^7^ Initially identified as a precursor state for blood cancers, CHIP is now established as a potent independent risk factor for a wide spectrum of non-malignant inflammatory diseases.

Mechanistically, this link is driven by the aberrant inflammatory activity of CHIP-mutant immune cells. Macrophages derived from hematopoietic cells with CHIP-driver mutations exhibit exaggerated pro-inflammatory responses, including heightened activation of the NLRP3 inflammasome and increased secretion of cytokines like interleukin-1β.^9, 10^ This creates a state of heightened systemic inflammation that is a causal driver of atherosclerotic cardiovascular disease and is also associated with chronic obstructive pulmonary disease, chronic kidney disease, and metabolic dysfunction-associated steatotic liver disease.^11–14^

Given the established role of inflammation in the pathogenesis of major eye diseases, including neuroinflammation in glaucoma and innate immune activation in AMD and DR, it is plausible that the systemic inflammation driven by CHIP may also contribute to the risk of these ocular conditions.^15, 16^ To date, however, the association between CHIP and major age-related eye diseases remains largely unexplored in large-scale prospective studies. Therefore, this study aimed to investigate the association of CHIP with the risk of four major age-related eye diseases in the UK Biobank and to explore the effects of CHIP clone size, driver gene identity, and potential inflammatory mediation.

## Methods

### Study Design and Population

This prospective cohort study was conducted using data from the UK Biobank, a large-scale, population-based cohort that enrolled more than 500,000 participants aged 40 to 69 years from 22 assessment centers across the United Kingdom between 2006 and 2010.^17^ At the baseline visit, participants provided detailed information via touchscreen questionnaires and verbal interviews, underwent physical measurements, and supplied biological samples for long-term storage and analysis. The UK Biobank study received approval from the North West Multi-centre Research Ethics Committee (reference 11/NW/0382), and all participants provided written informed consent. The analyses presented herein were performed under UK Biobank application number 101633. This study follows the Strengthening the Reporting of Observational Studies in Epidemiology (STROBE) reporting guideline.^18^

From the initial 501,936 participants, we excluded those lacking whole-exome sequencing data required for CHIP ascertainment (n = 32,886). Participants with prevalent diagnoses of the four age-related eye diseases of interest (cataract, glaucoma, AMD, and DR) at or before baseline were further excluded in outcome-specific analyses. This yielded a final analytical cohort of 436,469 participants for the incident disease analyses (**Figure 1**).

**Figure 1.**
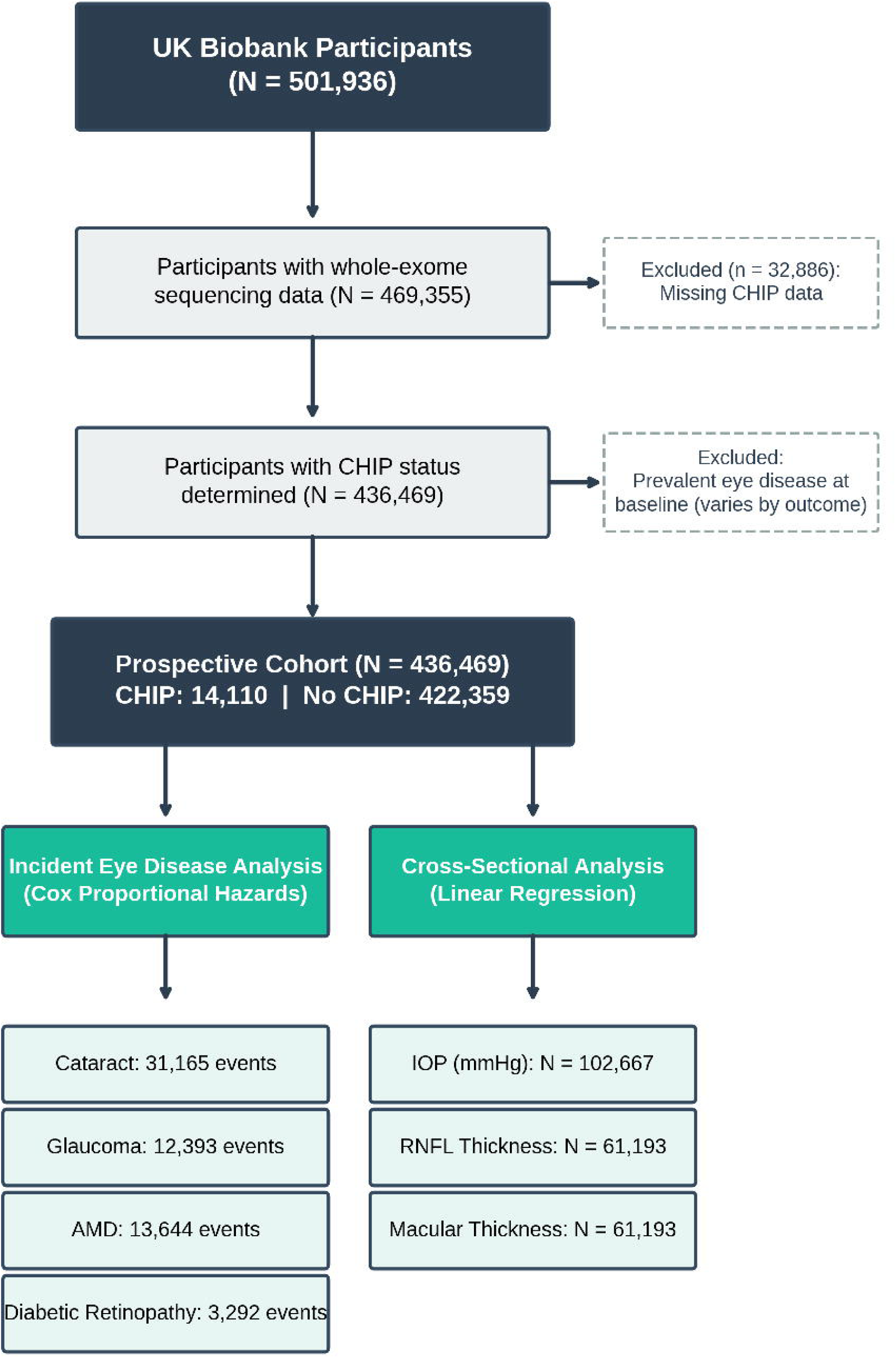
Flowchart of Participant Selection. Flowchart illustrating the selection of the study cohort from the UK Biobank. From an initial 501,936 participants, individuals were excluded due to the absence of whole-exome sequencing data or prevalent diagnoses of the four age-related eye diseases at baseline, resulting in a final analytical cohort of 436,469 participants for the incident disease analyses.

### Ascertainment of Clonal Hematopoiesis

CHIP status was determined using curated somatic mutation data derived from whole-exome sequencing of blood DNA, provided by the UK Biobank (Category 170; Data-Fields 30105-30107). The bioinformatic pipeline for variant calling has been described in detail by Vlasschaert et al.^19^ In brief, somatic mutations were identified using the Mutect2 algorithm from the Genome Analysis Toolkit in tumor-only mode, targeting a panel of 74 genes recurrently mutated in myeloid malignancies. A multi-step filtering process that integrates sequencing quality metrics, variant annotation, and population-level allele frequency data was applied to distinguish true somatic variants from germline variants and sequencing artifacts. CHIP was defined by the presence of at least one qualifying somatic mutation with a variant allele fraction (VAF) of 2% or greater.^7, 20^ CHIP carriers were further categorized by clone size into Small CHIP (2% ≤ VAF < 10%) and Large CHIP (VAF ≥ 10%), consistent with prior literature.^7^ Gene-specific analyses were performed for the three most commonly mutated driver genes (DNMT3A, TET2, and ASXL1), as well as a composite category encompassing all other less frequent driver mutations ("Other").

### Definition of Outcomes and Ocular Structural Measures

Incident eye disease outcomes were ascertained through algorithmic linkage to national electronic health records, including Hospital Episode Statistics (HES) for England, Scottish Morbidity Records (SMR), and the Patient Episode Database for Wales (PEDW). The follow-up period for each participant commenced at the date of baseline assessment and extended until the date of first diagnosis of the outcome of interest, death, loss to follow-up, or the administrative censoring date (March 31, 2022), whichever occurred first. Cataract was identified using ICD-10 codes H25, H26, and H28.0, as well as OPCS-4 procedure codes C71-C75; glaucoma using ICD-10 codes H40 and H42; AMD using ICD-10 code H35.3; and DR using ICD-10 codes H36.0, E10.3, E11.3, E13.3, and E14.3 (**eTable 1**).

Cross-sectional ocular structural measures were obtained from the enhanced ophthalmic assessment conducted on a subset of UK Biobank participants.^21^ Intraocular pressure (IOP) was measured using the Ocular Response Analyzer (Reichert Technologies), which provides corneal-compensated IOP (IOPcc); this metric was used for our analyses as it is less influenced by corneal biomechanical properties.^22^ Retinal nerve fiber layer (RNFL) thickness and central macular thickness were measured using spectral-domain optical coherence tomography (Topcon 3D OCT 1000 Mk2), with automated segmentation and quality control as previously described.^23^

### Assessment of Covariates

Baseline covariates were collected during the initial assessment visit. Sociodemographic variables included age at recruitment, sex, and self-reported ethnicity. Lifestyle and socioeconomic factors included educational attainment, Townsend deprivation index, smoking status, alcohol consumption frequency, physical activity level (International Physical Activity Questionnaire categories), sleep duration, and body mass index (BMI). Prevalent clinical comorbidities, including type 2 diabetes, hypertension, and hyperlipidemia, were ascertained from a combination of self-report, medication records, and linked hospital inpatient diagnoses (**eTable 1**).

### Statistical Analysis

Baseline characteristics were summarized as means (standard deviations) for continuous variables and frequencies (percentages) for categorical variables. Between-group differences were assessed using independent-samples t-tests for continuous variables and chi-squared tests for categorical variables.

Multivariable Cox proportional hazards regression models were used to estimate hazard ratios (HRs) and 95% confidence intervals (CIs) for the associations of CHIP status (overall, by clone size, and by driver gene) with the risk of each incident eye disease. Three nested models with sequential covariate adjustment were constructed: Model 1 adjusted for age, sex, and ethnicity; Model 2 additionally adjusted for education level, Townsend deprivation index, smoking status, alcohol frequency, BMI, physical activity, and sleep duration; Model 3 (the primary, fully adjusted model) further adjusted for prevalent diabetes, hypertension, and hyperlipidemia. The proportional hazards assumption was verified using Schoenfeld residuals.

Cross-sectional associations between CHIP and continuous ocular structural measures (IOPcc, RNFL thickness, and macular thickness) were assessed using multivariable linear regression with the same three levels of covariate adjustment. To characterize the dose-response relationship between CHIP clone size and incident eye disease risk, we modeled VAF as a continuous variable using restricted cubic splines (RCS) with four knots placed at the 5th, 35th, 65th, and 95th percentiles of the VAF distribution among CHIP carriers. Non-linearity was assessed by a likelihood ratio test comparing the spline model to a model with only a linear term. Pre-specified subgroup analyses were conducted to evaluate potential effect modification by age (<60 vs ≥60 years), sex (female vs male), BMI (<30 vs ≥30 kg/m²), prevalent diabetes (no vs yes), and prevalent hypertension (no vs yes). P-values for interaction were derived from likelihood ratio tests comparing models with and without multiplicative interaction terms.

To explore potential biological pathways underlying the observed associations, we performed mediation analyses using the product-of-coefficients approach.^24^ Candidate mediators included hematologic and inflammatory biomarkers: white blood cell count, neutrophil count, monocyte count, lymphocyte count, platelet count, and C-reactive protein (CRP). The proportion of the total effect mediated through each biomarker was estimated with 95% CIs derived from 1000 bootstrap resamples.

As sensitivity analyses, we repeated the primary analyses after (1) excluding events occurring within the first 2 years of follow-up (landmark analysis) to minimize the possibility of reverse causation, and (2) restricting the cohort to participants of White European ancestry to assess the robustness of findings across population subgroups.

All statistical analyses were performed using R software, version 4.4.0 (R Foundation for Statistical Computing). A two-sided P value of less than .05 was considered statistically significant.

## Results

### Baseline Characteristics of the Study Population

Of the 436,469 participants included in the analytical cohort, 14,110 (3.2%) were identified as having CHIP. The mean (SD) age of the cohort was 56.4 (8.1) years, and 237,745 (54.5%) were women. Compared with participants without CHIP, those with CHIP were older (mean age, 60.5 vs 56.2 years; P < .001), more likely to be former smokers, and had a higher prevalence of cardiometabolic comorbidities, including hypertension (58.1% vs 50.4%; P < .001), diabetes (6.8% vs 5.4%; P < .001), and hyperlipidemia (13.5% vs 10.0%; P < .001). Individuals with CHIP also exhibited higher levels of systemic inflammation, as indicated by elevated white blood cell counts and C-reactive protein levels (**Table 1**).

**Table 1.**
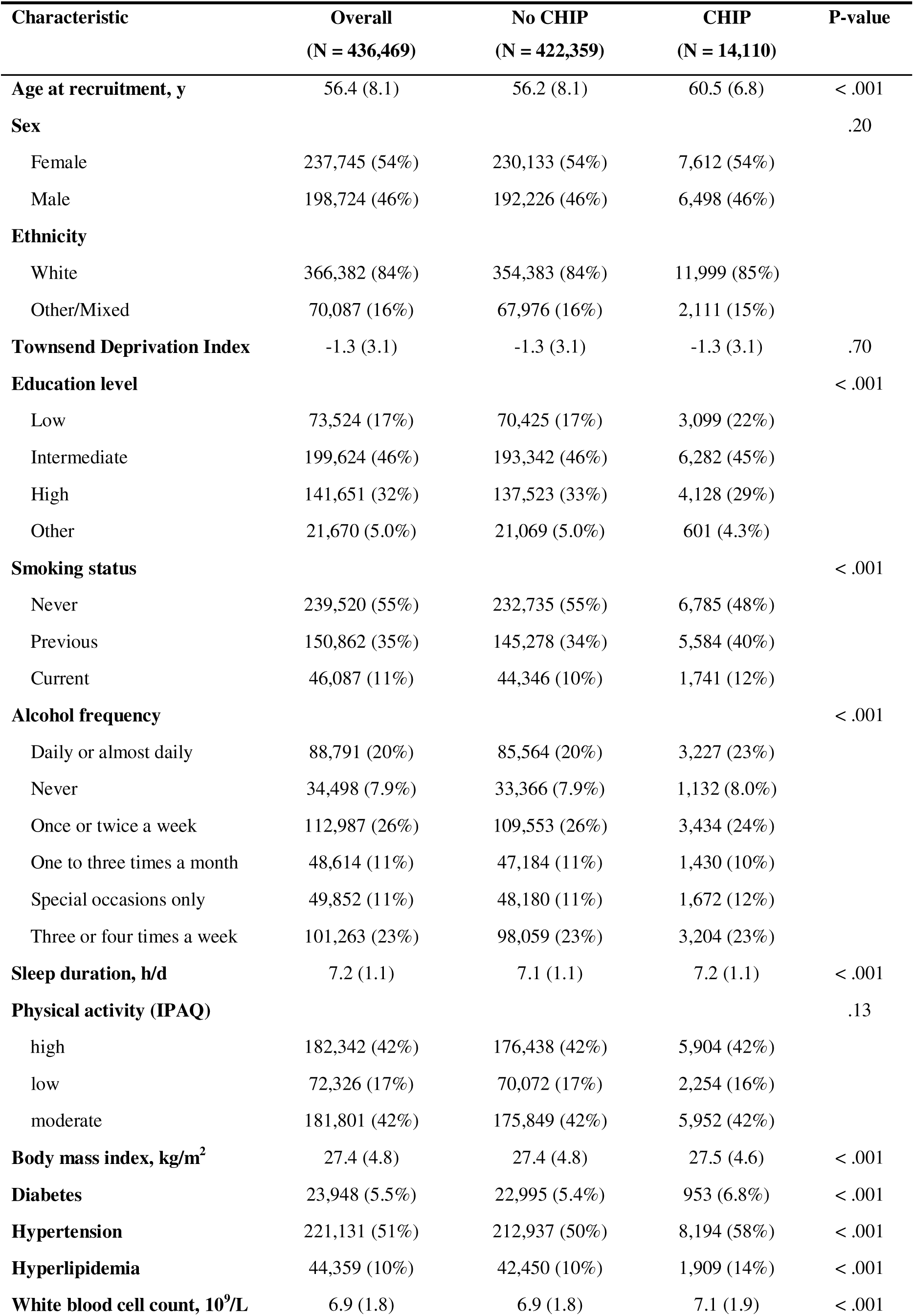
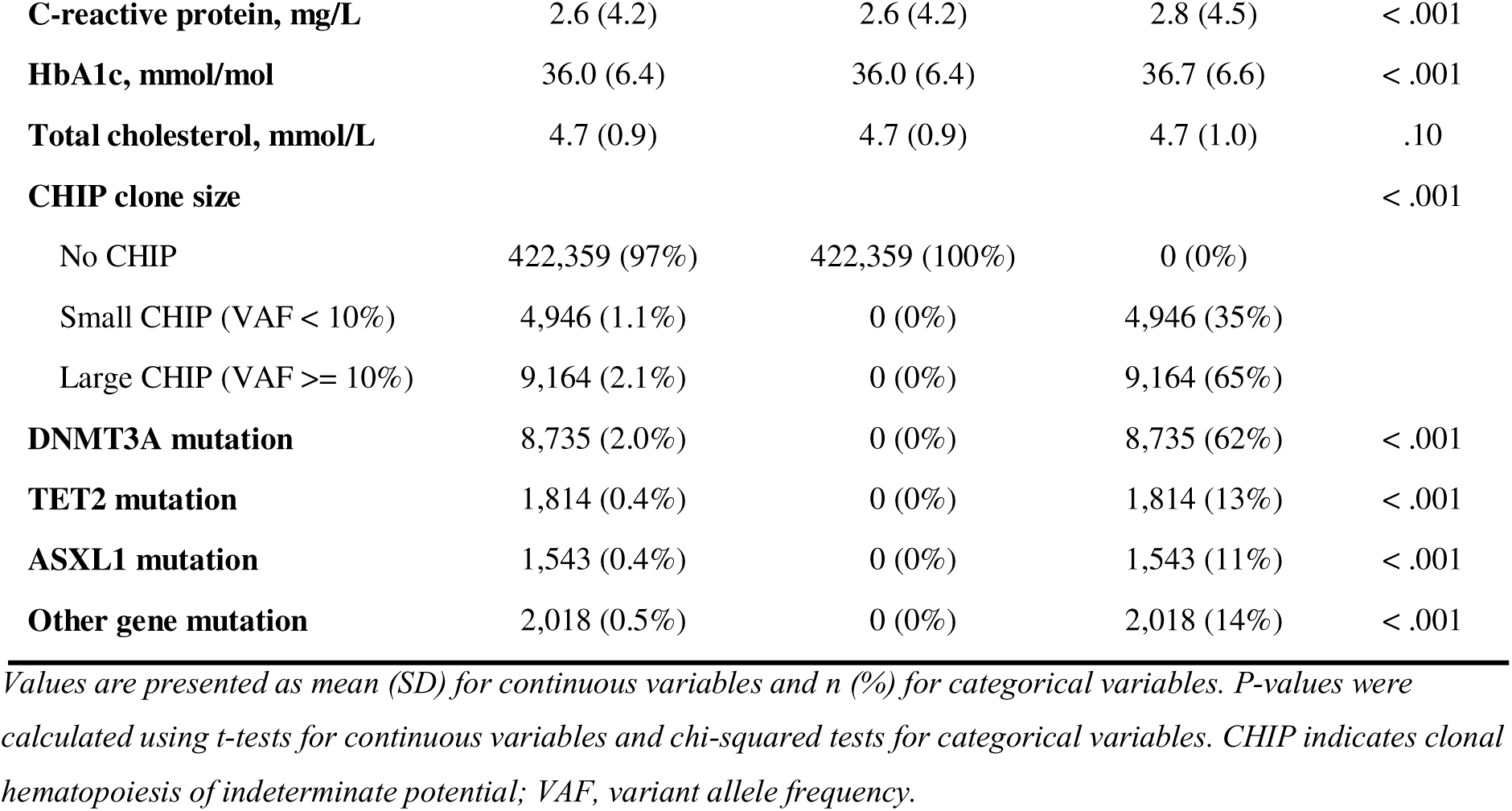
Baseline Characteristics of Study Participants by CHIP Status.

| **Characteristic** | **Overall**<br>**(N = 436,469)** | **No CHIP**<br>**(N = 422,359)** | **CHIP**<br>**(N = 14,110)** | **P-value** |
| --- | --- | --- | --- | --- |
| **Age at recruitment, y** | 56.4 (8.1) | 56.2 (8.1) | 60.5 (6.8) | < .001 |
| **Sex** |  |  |  | .20 |
| Female | 237,745 (54%) | 230,133 (54%) | 7,612 (54%) |  |
| Male | 198,724 (46%) | 192,226 (46%) | 6,498 (46%) |  |
| **Ethnicity** |  |  |  |  |
| White | 366,382 (84%) | 354,383 (84%) | 11,999 (85%) |  |
| Other/Mixed | 70,087 (16%) | 67,976 (16%) | 2,111 (15%) |  |
| **Townsend Deprivation Index** | -1.3 (3.1) | -1.3 (3.1) | -1.3 (3.1) | .70 |
| **Education level** |  |  |  | < .001 |
| Low | 73,524 (17%) | 70,425 (17%) | 3,099 (22%) |  |
| Intermediate | 199,624 (46%) | 193,342 (46%) | 6,282 (45%) |  |
| High | 141,651 (32%) | 137,523 (33%) | 4,128 (29%) |  |
| Other | 21,670 (5.0%) | 21,069 (5.0%) | 601 (4.3%) |  |
| **Smoking status** |  |  |  | < .001 |
| Never | 239,520 (55%) | 232,735 (55%) | 6,785 (48%) |  |
| Previous | 150,862 (35%) | 145,278 (34%) | 5,584 (40%) |  |
| Current | 46,087 (11%) | 44,346 (10%) | 1,741 (12%) |  |
| **Alcohol frequency** |  |  |  | < .001 |
| Daily or almost daily | 88,791 (20%) | 85,564 (20%) | 3,227 (23%) |  |
| Never | 34,498 (7.9%) | 33,366 (7.9%) | 1,132 (8.0%) |  |
| Once or twice a week | 112,987 (26%) | 109,553 (26%) | 3,434 (24%) |  |
| One to three times a month | 48,614 (11%) | 47,184 (11%) | 1,430 (10%) |  |
| Special occasions only | 49,852 (11%) | 48,180 (11%) | 1,672 (12%) |  |
| Three or four times a week | 101,263 (23%) | 98,059 (23%) | 3,204 (23%) |  |
| **Sleep duration, h/d** | 7.2 (1.1) | 7.1 (1.1) | 7.2 (1.1) | < .001 |
| **Physical activity (IPAQ)** |  |  |  | .13 |
| high | 182,342 (42%) | 176,438 (42%) | 5,904 (42%) |  |
| low | 72,326 (17%) | 70,072 (17%) | 2,254 (16%) |  |
| moderate | 181,801 (42%) | 175,849 (42%) | 5,952 (42%) |  |
| **Body mass index, kg/m<sup>2</sup>** | 27.4 (4.8) | 27.4 (4.8) | 27.5 (4.6) | < .001 |
| **Diabetes** | 23,948 (5.5%) | 22,995 (5.4%) | 953 (6.8%) | < .001 |
| **Hypertension** | 221,131 (51%) | 212,937 (50%) | 8,194 (58%) | < .001 |
| **Hyperlipidemia** | 44,359 (10%) | 42,450 (10%) | 1,909 (14%) | < .001 |
| **White blood cell count, 10<sup>9</sup>/L** | 6.9 (1.8) | 6.9 (1.8) | 7.1 (1.9) | < .001 |
| **C-reactive protein, mg/L** | 2.6 (4.2) | 2.6 (4.2) | 2.8 (4.5) | < .001 |
| **HbA1c, mmol/mol** | 36.0 (6.4) | 36.0 (6.4) | 36.7 (6.6) | < .001 |
| **Total cholesterol, mmol/L** | 4.7 (0.9) | 4.7 (0.9) | 4.7 (1.0) | .10 |
| **CHIP clone size** |  |  |  | < .001 |
| No CHIP | 422,359 (97%) | 422,359 (100%) | 0 (0%) |  |
| Small CHIP (VAF < 10%) | 4,946 (1.1%) | 0 (0%) | 4,946 (35%) |  |
| Large CHIP (VAF >= 10%) | 9,164 (2.1%) | 0 (0%) | 9,164 (65%) |  |
| **DNMT3A mutation** | 8,735 (2.0%) | 0 (0%) | 8,735 (62%) | < .001 |
| **TET2 mutation** | 1,814 (0.4%) | 0 (0%) | 1,814 (13%) | < .001 |
| **ASXL1 mutation** | 1,543 (0.4%) | 0 (0%) | 1,543 (11%) | < .001 |
| **Other gene mutation** | 2,018 (0.5%) | 0 (0%) | 2,018 (14%) | < .001 |

Values are presented as mean (SD) for continuous variables and n (%) for categorical variables. P-values were calculated using t-tests for continuous variables and chi-squared tests for categorical variables. CHIP indicates clonal hematopoiesis of indeterminate potential; VAF, variant allele frequency.

Among participants with CHIP, DNMT3A was the most commonly mutated gene (62.0%), followed by TET2 (12.9%), ASXL1 (11.0%), and other driver genes (14.3%). A total of 9,164 individuals (65.0% of CHIP carriers) were classified as having large CHIP clones (VAF ≥10%).

### Association of CHIP with Incident Eye Diseases

Over a median follow-up of 13.1 years, we documented 31,165 incident cases of cataract, 12,393 cases of glaucoma, 13,644 cases of AMD, and 3,292 cases of DR. In fully adjusted Cox proportional hazards models (Model 3), the presence of any CHIP was significantly associated with an increased risk of developing cataract (HR, 1.08; 95% CI, 1.03-1.14; P = .003), AMD (HR, 1.12; 95% CI, 1.04-1.21; P = .004), and DR (HR, 1.41; 95% CI, 1.20-1.64; P < .001). The association with glaucoma was not statistically significant (HR, 1.08; 95% CI, 0.99-1.17; P = .095) (**Figure 2**; **eTable 2**). The associations remained consistent in sensitivity analyses, including a 2-year landmark analysis and when restricting the cohort to participants of White European ancestry (**eTable 3**).

**Figure 2.**
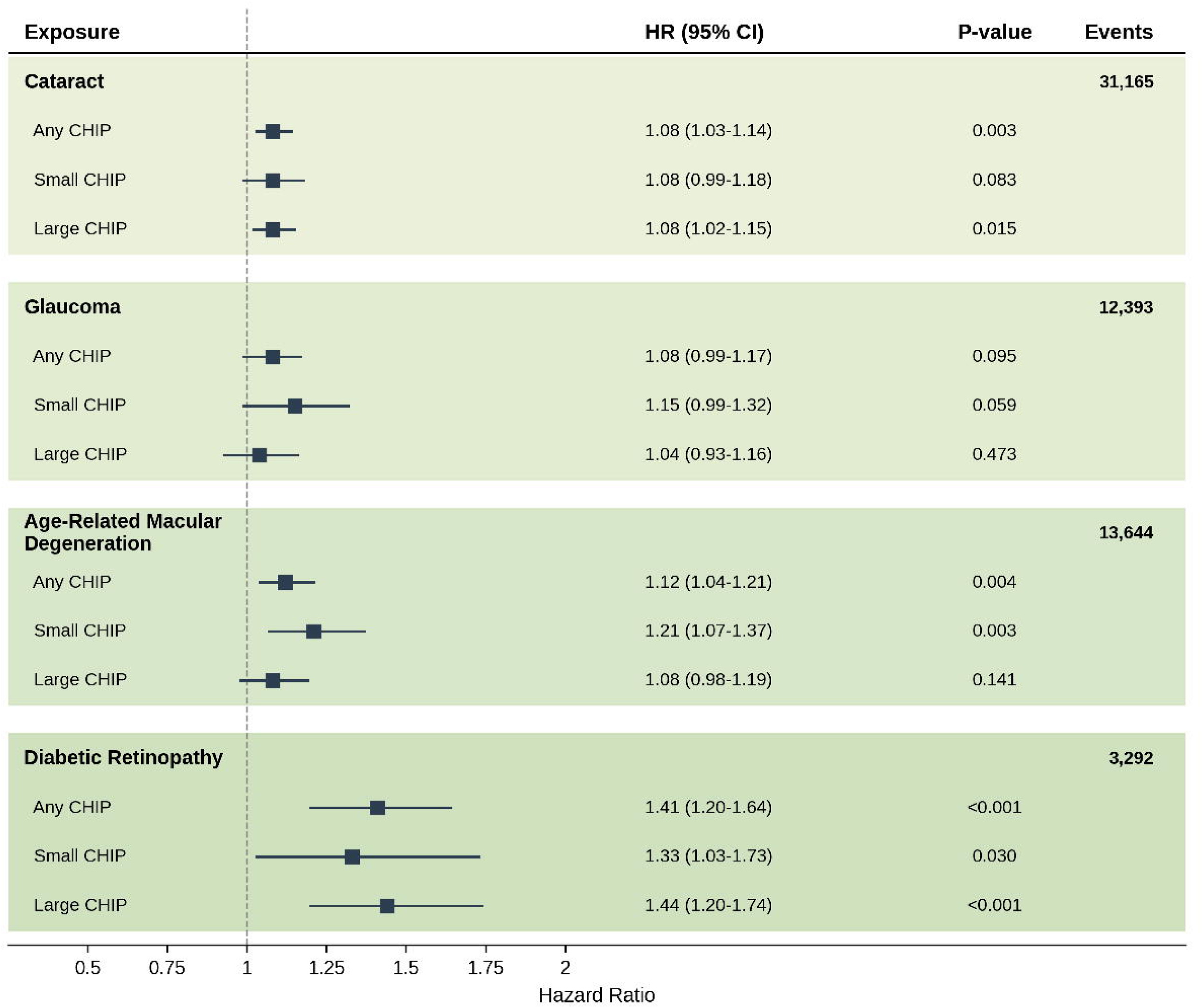
Association of Clonal Hematopoiesis of Indeterminate Potential (CHIP) with Incident Eye Diseases. Forest plot showing hazard ratios (HRs) and 95% CIs for the associations of CHIP, stratified by presence (Any CHIP) and clone size (Small CHIP: 2% ≤ Variant Allele Fraction [VAF] < 10%; Large CHIP: VAF ≥10%), with incident cataract, glaucoma, age-related macular degeneration (AMD), and diabetic retinopathy (DR). Results are from the fully adjusted Cox proportional hazards model (Model 3), adjusted for age, sex, ethnicity, education, Townsend deprivation index, smoking status, alcohol frequency, body mass index, physical activity, sleep duration, prevalent diabetes, hypertension, and hyperlipidemia.

When stratified by clone size, large CHIP (VAF ≥10%) was the primary driver of the association with cataract (HR, 1.08; 95% CI, 1.02-1.15) and DR (HR, 1.44; 95% CI, 1.20-1.74). Conversely, the risk of AMD was predominantly associated with small CHIP (VAF <10%) (HR, 1.21; 95% CI, 1.07-1.37) (Figure 2; eTable 2). In gene-specific analyses, mutations in the "Other" gene category were associated with the highest risk for AMD (HR, 1.23; 95% CI, 1.01-1.50) and DR (HR, 1.76; 95% CI, 1.27-2.45). DNMT3A mutations were also significantly associated with an increased risk of DR (HR, 1.36; 95% CI, 1.10-1.66) (**eFigure 1**; **eTable 4**).

RCS analyses showed a generally linear dose-response relationship between VAF and the risk of cataract, glaucoma, and AMD (P for nonlinearity = .21, .50, and .89, respectively). However, a significant nonlinear, J-shaped association was observed for DR, with risk increasing more steeply at higher VAFs (P for nonlinearity < .001) (**Figure 3**).

**Figure 3.**
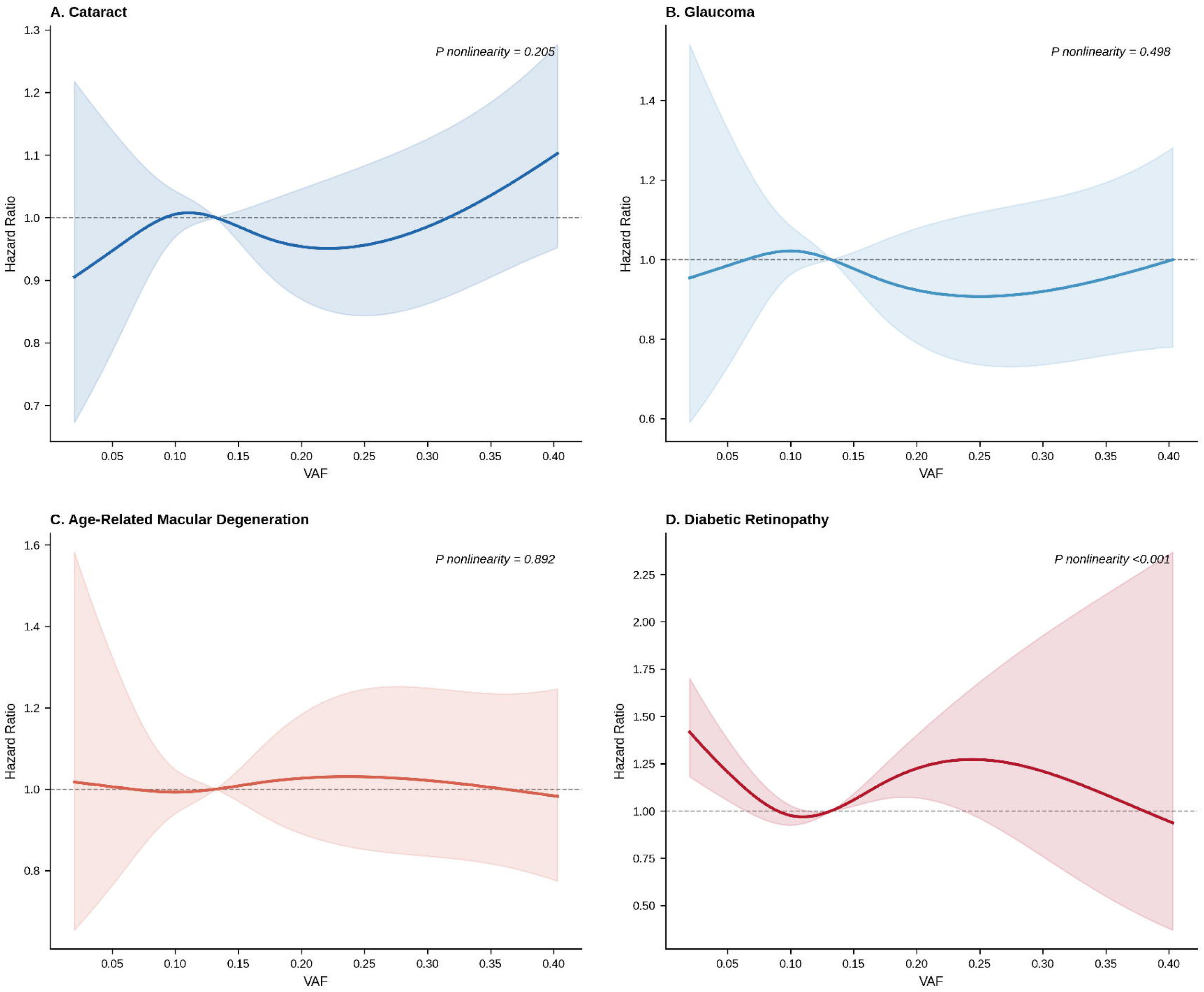
Dose-Response Relationship Between CHIP Clone Size and Risk of Incident Eye Diseases. Restricted cubic spline plots illustrating the dose-response relationship between CHIP variant allele fraction (VAF) as a continuous variable and the risk of incident cataract, glaucoma, age-related macular degeneration (AMD), and diabetic retinopathy (DR). The solid lines represent the estimated hazard ratios, and the shaded areas represent the 95% confidence intervals. Splines were modeled with four knots. P-values for nonlinearity are shown. Models were fully adjusted as described in Figure 2.

### Subgroup Analyses

Subgroup analyses revealed significant interactions (**eFigure 2**; **eTable 5**). The association between CHIP and cataract was stronger in younger individuals (age <60 years: HR, 1.48 vs ≥60 years: HR, 1.13; P for interaction < .001). For glaucoma, the association was significant only among individuals without hypertension (HR, 1.22; 95% CI, 1.06-1.40) but not in those with hypertension (HR, 1.00; 95% CI, 0.89-1.12), with a significant interaction (P for interaction = .012).

### Mediation Analyses

Mediation analyses suggested that systemic inflammatory and hematologic biomarkers partially mediated the observed associations. For cataract, the proportion of the effect mediated was highest for neutrophil count (3.57%) and white blood cell count (2.66%). For DR, neutrophil count also appeared to be a key mediator, accounting for 4.68% of the total effect (**Figure 4**).

**Figure 4.**
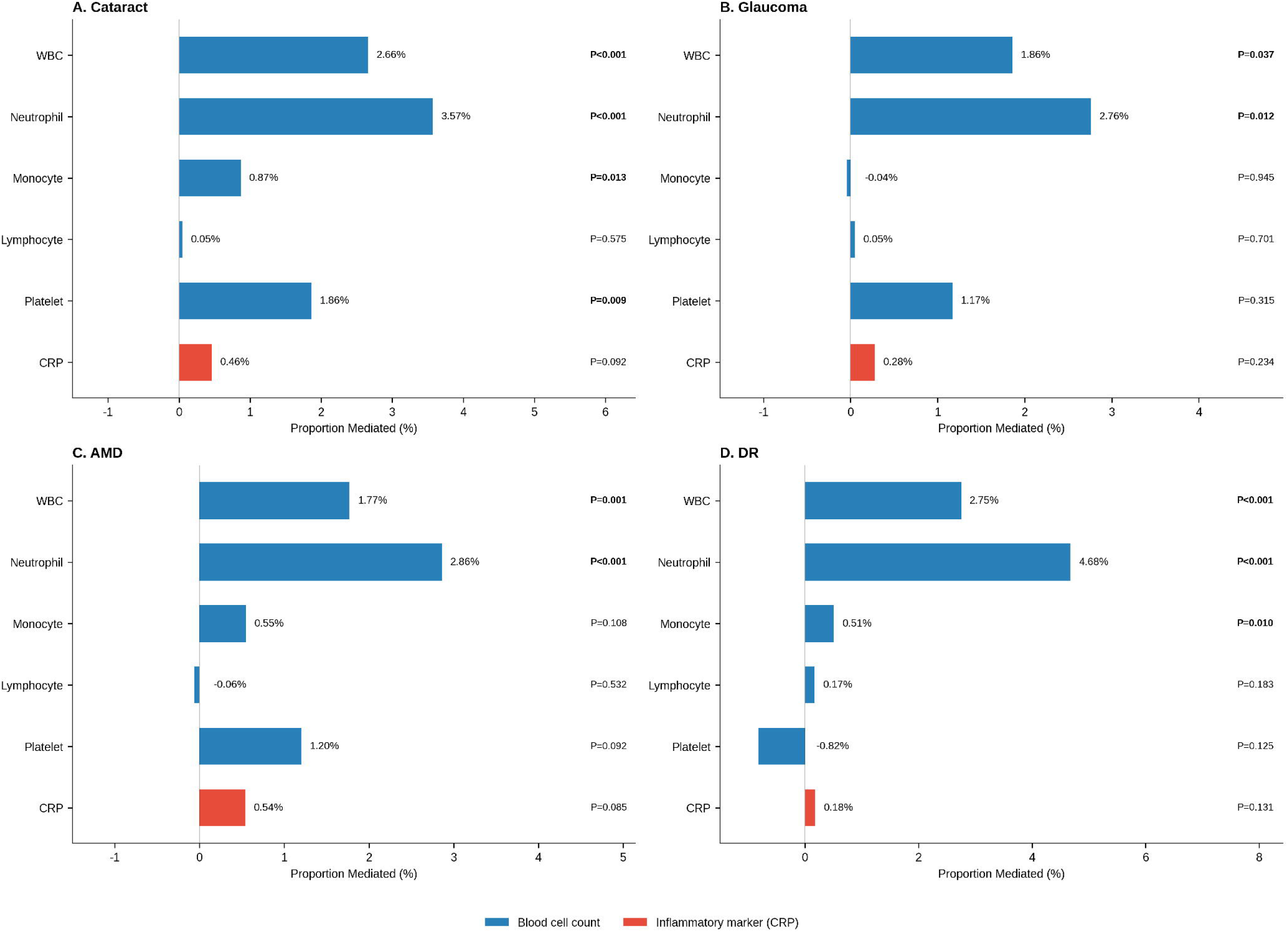
Mediation of the Association Between CHIP and Incident Eye Diseases by Inflammatory Biomarkers. Bar plot showing the proportion of the total effect of CHIP on incident eye diseases mediated by candidate inflammatory and hematologic biomarkers. Results are shown for all four outcomes. The proportion mediated and 95% CIs were estimated using a mediation framework with 1000 bootstrap resamples. Significant mediation pathways (P < .05) are indicated in bold.

### Association of CHIP with Ocular Structural Measures

In cross-sectional analyses of ocular structural parameters, we found no significant association between the presence of any CHIP and corneal-compensated IOP, mean RNFL thickness, or macular thickness in fully adjusted models (**eTable 6**). However, in gene-specific analyses, the presence of a TET2 mutation was significantly associated with lower IOP (β, −0.39 mmHg; 95% CI, −0.73 to −0.05; P = .027) (**eTable 7**).

## Discussion

In this prospective study of 436,469 UK Biobank participants, CHIP was significantly associated with an increased risk of cataract, AMD, and DR, whereas no significant association was observed for glaucoma. The magnitude of risk varied by eye disease, CHIP clone size, and driver gene mutation. These associations were partially mediated by systemic inflammation, particularly neutrophil count, and remained robust in sensitivity analyses. Our findings position CHIP as a novel, independent risk factor for several major age-related eye diseases, linking systemic somatic mutations to ocular pathology through inflammatory pathways.

To our knowledge, this study provides the most comprehensive investigation to date of the association between CHIP and a broad spectrum of age-related eye diseases in a large-scale prospective setting. The findings substantially expand upon the limited existing literature. A recent conference abstract reported an association between CHIP and AMD in the UK Biobank, which is consistent with our results.^25^ Our analysis provides more granular detail, notably identifying that the AMD risk was predominantly driven by smaller clones (VAF <10%), suggesting that even early-stage clonal expansion may be sufficient to trigger or accelerate AMD-related pathology.

Furthermore, our strong association between CHIP and incident DR (HR, 1.41) aligns with a recent study that also found CHIP to be a risk factor for diabetic microvascular complications, including DR, in a cohort of individuals with type 2 diabetes.^26^ The associations we observed with cataract and the suggestive association with glaucoma are, to our knowledge, entirely novel and have not been previously reported.

The observed associations are biologically plausible and likely mediated by the pro-inflammatory state induced by CHIP. The central role of inflammation in the pathogenesis of AMD and DR is well-established.^27, 28^ In AMD, chronic activation of the complement system and infiltration of macrophages in the retinal pigment epithelium (RPE) and Bruch’s membrane contribute to drusen formation and disease progression.^29^ For DR, hyperglycemia-induced inflammation promotes retinal leukostasis, microvascular damage, and breakdown of the blood-retinal barrier, with neutrophils playing a key role.^30^ Our mediation analysis, which identified neutrophil count as the most significant mediator for the associations with both DR and cataract, provides strong human epidemiological evidence supporting this inflammatory link. The established mechanism for CHIP-driven systemic inflammation involves the hyperactivation of the NLRP3 inflammasome in mutant hematopoietic cells (particularly TET2-mutant macrophages), leading to increased production of pro-inflammatory cytokines such as interleukin-1β (IL-1β) and IL-6.^9, 31^ This same NLRP3 inflammasome pathway has been independently implicated in the pathogenesis of AMD, glaucoma, and cataract, providing a compelling mechanistic convergence.^32, 33^

Our study revealed distinct patterns of association for different CHIP characteristics. The J-shaped dose-response curve for DR, with risk accelerating at higher VAFs, suggests a threshold effect, where larger, more established clones exert a more potent inflammatory effect on the susceptible retinal microvasculature. Conversely, the finding that small CHIP clones were the primary drivers of AMD risk is intriguing. It may be that the chronic, low-grade inflammation initiated by smaller clones is sufficient to dysregulate the delicate immune balance at the RPE-Bruch’s membrane interface, a key initiating step in AMD. The gene-specific findings, such as the strong association of the "Other" gene category (including spliceosome mutations) with DR and AMD, and the paradoxical association of TET2 mutations with lower IOP, underscore the biological heterogeneity of CHIP. The TET2-IOP finding, while unexpected, could relate to the known role of TET enzymes in regulating aqueous humor outflow dynamics in the trabecular meshwork, suggesting a complex, potentially IOP-independent neuroinflammatory mechanism in glaucoma.^34^

The significant interaction between CHIP and age for cataract risk, with a much stronger effect in individuals younger than 60, is a notable finding. This suggests that in younger individuals, CHIP may be a more prominent contributor to cataractogenesis, while in older individuals, its association may be overshadowed by other cumulative age-related factors like oxidative stress.^35^ Similarly, the significant association of CHIP with glaucoma only in participants without hypertension suggests that in the absence of a strong competing risk factor like hypertensive vascular damage, the pro-inflammatory effect of CHIP on the optic nerve head becomes more apparent. This highlights the complex interplay between genetic predisposition (CHIP) and traditional risk factors in the pathogenesis of eye disease.

Our findings have significant clinical implications. They identify CHIP as a novel, non-ocular risk factor for several of the most common causes of blindness worldwide. This raises the possibility that individuals with known CHIP, who are increasingly being identified in clinical practice, may benefit from more vigilant ophthalmic screening for early detection of eye disease. Furthermore, our results provide a rationale for exploring anti-inflammatory therapies as a potential strategy to mitigate the ocular risks associated with CHIP. The success of the CANTOS trial, which demonstrated that inhibiting IL-1β could reduce cardiovascular events in individuals with residual inflammatory risk, provides a powerful proof-of-concept.^36^ Whether similar targeted anti-inflammatory strategies could prevent or slow the progression of CHIP-associated eye diseases is a promising avenue for future clinical investigation.

This study has several major strengths, including its large sample size, prospective design with over a decade of follow-up, and the use of centrally adjudicated, exome-wide CHIP data. The comprehensive analysis of four distinct eye diseases, along with detailed gene- and clone size-specific analyses, provides a uniquely granular perspective. However, our study also has limitations. First, as an observational study, we cannot definitively establish causality, although the dose-response relationships and consistency with known biological mechanisms provide supportive evidence. Second, the diagnoses of eye diseases were based on ICD codes from electronic health records, which may be subject to misclassification, although this is a widely accepted method in large-scale epidemiology. Third, the UK Biobank cohort is known to have a "healthy volunteer" selection bias and is predominantly of European ancestry, which may limit the generalizability of our findings to other populations.^37^ Finally, while our study is the largest of its kind, the number of events for some analyses, particularly for rarer gene mutations, was limited.

## Conclusions

In conclusion, our findings from this large, prospective study demonstrate that CHIP is a significant and independent risk factor for the development of cataract, AMD, and DR. The risk is heterogeneous, varying by the specific eye disease, CHIP clone size, and driver gene. These associations appear to be at least partially mediated by systemic inflammation. These findings establish a novel link between somatic mutations in the hematopoietic system and the pathogenesis of major age-related eye diseases, opening new avenues for risk stratification and the development of targeted preventive therapies.

## Additional Sections

### Author Contributions

**Conceptualization:** Ruijie Xie, Ben Schöttker.

**Data Curation:** Ruijie Xie.

**Formal Analysis:** Ruijie Xie.

**Funding Acquisition:** Ben Schöttker.

**Methodology:** Ruijie Xie, Ben Schöttker.

**Project Administration:** Ben Schöttker.

**Supervision:** Ben Schöttker.

**Visualization:** Ruijie Xie.

**Writing - Original Draft:** Ruijie Xie.

**Writing - Review & Editing:** Ruijie Xie, Ben Schöttker.

### Conflict of Interest Disclosures

None reported.

### Funding/Support

This research was conducted using the UK Biobank Resource under Application Number 101633.

### Role of the Funder/Sponsor

The funders had no role in the design and conduct of the study; collection, management, analysis, and interpretation of the data; preparation, review, or approval of the manuscript; and decision to submit the manuscript for publication.

### Data Availability Statement

Data from the UK Biobank are available to bona fide researchers upon application through the UK Biobank Access Management System (https://www.ukbiobank.ac.uk/enable-your-research/apply-for-access). The analytical code supporting the findings of this study is available from the corresponding author upon reasonable request.

## Abbreviations

AMD: Age-Related Macular Degeneration
ASXL1: ASXL Transcriptional Regulator 1
BMI: Body Mass Index
CHIP: Clonal Hematopoiesis of Indeterminate Potential
CI: Confidence Interval
CRP: C-Reactive Protein
DNMT3A: DNA Methyltransferase 3 Alpha
DR: Diabetic Retinopathy
HES: Hospital Episode Statistics
HR: Hazard Ratio
ICD-10: International Classification of Diseases, 10th Revision
IL: Interleukin
IOP: Intraocular Pressure
NLRP3: NLR Family Pyrin Domain Containing 3
OPCS-4: Office of Population Censuses and Surveys Classification of Interventions and Procedures, 4th Revision
PEDW: Patient Episode Database for Wales
RCS: Restricted Cubic Spline
RNFL: Retinal Nerve Fiber Layer
SD: Standard Deviation
SMR: Scottish Morbidity Records
TET2: Tet Methylcytosine Dioxygenase 2
VAF: Variant Allele Fraction

## Notes

### Competing Interest Statement

The authors have declared no competing interest.

### Funding Statement

This research was conducted using the UK Biobank Resource under Application Number 101633. The UK Biobank was established by the Wellcome Trust, Medical Research Council, Department of Health, Scottish Government, Northwest Regional Development Agency, Welsh Assembly Government, and the British Heart Foundation. The funders had no role in the study design, data collection, data analysis, interpretation, or the decision to submit the manuscript for publication. The authors received no specific funding for this work.

### Author Declarations

The North West Multi-centre Research Ethics Committee gave ethical approval for this work (reference 11/NW/0382). All UK Biobank participants provided written informed consent at enrollment. This research was conducted using the UK Biobank Resource under Application Number 101633.

