## Supplementary Material for "Clonal Hematopoiesis of Indeterminate Potential and Risk of Major Age-Related Eye Diseases"

**eTable 1. Variable Definitions and ICD-10 Codes**

| **Variable** | **Definition / ICD-10 Code** | **Data Source** |
| --- | --- | --- |
| **Outcomes** |  |  |
| Cataract | ICD-10: H25, H26, H28.0; OPCS-4: C71-C75 | Hospital inpatient records |
| Glaucoma | ICD-10: H40, H42 | Hospital inpatient records |
| Age-related macular degeneration | ICD-10: H35.3 | Hospital inpatient records |
| Diabetic retinopathy | ICD-10: H36.0, E10.3, E11.3, E13.3, E14.3 | Hospital inpatient records |
| **Exposures** |  |  |
| CHIP status | Presence of somatic mutation (VAF ≥ 2%) in CHIP driver gene | Whole exome sequencing |
| CHIP clone size | Small: VAF < 10%; Large: VAF ≥ 10% | Whole exome sequencing |
| CHIP gene | DNMT3A, TET2, ASXL1, Other | Whole exome sequencing |
| **Structural Measures** |  |  |
| Intraocular pressure (IOP) | Corneal-compensated IOP, mmHg | Ocular assessment |
| RNFL thickness | Retinal nerve fiber layer thickness, μm | Optical coherence tomography |
| Macular thickness | Central macular thickness, μm | Optical coherence tomography |
| **Covariates** |  |  |
| Age | Age at recruitment, years | Baseline assessment |
| Sex | Male / Female | Baseline assessment |
| Ethnicity | White / Non-White | Baseline assessment |
| Townsend Deprivation Index | Area-based deprivation score | Baseline assessment |
| Education level | Low / Intermediate / High / Others | Baseline assessment |
| Smoking status | Never / Previous / Current | Baseline assessment |
| Alcohol frequency | Daily or almost daily / 3-4 times/week / 1-2 times/week / Others | Baseline assessment |
| BMI | Body mass index, kg/m² | Baseline assessment |
| Physical activity | IPAQ categories: Low / Moderate / High | Baseline assessment |
| Sleep duration | Hours per day | Baseline assessment |
| Diabetes | Self-reported or ICD-10: E10-E14 | Baseline + Hospital records |
| Hypertension | Self-reported or ICD-10: I10-I15 | Baseline + Hospital records |
| Hyperlipidemia | Self-reported or ICD-10: E78 | Baseline + Hospital records |

**eTable 2. Association of CHIP With Incident Eye Diseases: Cox Proportional Hazards Regression**

| **Outcome** | **Exposure** | **Events** | **Model 1 HR (95% CI)** | **Model 2 HR (95% CI)** | **Model 3 HR (95% CI)** |
| --- | --- | --- | --- | --- | --- |
| **Cataract** | Any CHIP | 31,165 | 1.09 (1.03-1.15) (P = .001) | 1.08 (1.03-1.14) (P = .003) | 1.08 (1.03-1.14) (P = .003) |
|  | Small CHIP |  | 1.09 (1.00-1.19) (P = .061) | 1.08 (0.99-1.18) (P = .088) | 1.08 (0.99-1.18) (P = .083) |
|  | Large CHIP |  | 1.09 (1.02-1.16) (P = .007) | 1.08 (1.02-1.15) (P = .015) | 1.08 (1.02-1.15) (P = .015) |
| **Glaucoma** | Any CHIP | 12,393 | 1.08 (0.99-1.18) (P = .084) | 1.08 (0.99-1.17) (P = .093) | 1.08 (0.99-1.17) (P = .095) |
|  | Small CHIP |  | 1.15 (1.00-1.32) (P = .054) | 1.15 (1.00-1.32) (P = .057) | 1.15 (0.99-1.32) (P = .059) |
|  | Large CHIP |  | 1.04 (0.94-1.16) (P = .447) | 1.04 (0.93-1.16) (P = .468) | 1.04 (0.93-1.16) (P = .473) |
| **AMD** | Any CHIP | 13,644 | 1.13 (1.04-1.22) (P = .003) | 1.12 (1.04-1.21) (P = .004) | 1.12 (1.04-1.21) (P = .004) |
|  | Small CHIP |  | 1.22 (1.07-1.38) (P = .002) | 1.21 (1.07-1.38) (P = .003) | 1.21 (1.07-1.37) (P = .003) |
|  | Large CHIP |  | 1.08 (0.98-1.19) (P = .118) | 1.08 (0.97-1.19) (P = .147) | 1.08 (0.98-1.19) (P = .141) |
| **DR** | Any CHIP | 3,292 | 1.37 (1.18-1.60) (P < .001) | 1.37 (1.17-1.59) (P < .001) | 1.41 (1.20-1.64) (P < .001) |
|  | Small CHIP |  | 1.34 (1.03-1.73) (P = .029) | 1.30 (1.01-1.69) (P = .046) | 1.33 (1.03-1.73) (P = .030) |
|  | Large CHIP |  | 1.39 (1.15-1.68) (P < .001) | 1.40 (1.16-1.69) (P < .001) | 1.44 (1.20-1.74) (P < .001) |

*Model 1: adjusted for age, sex, and ethnicity. Model 2: Model 1 + ethnicity, Townsend Deprivation Index, education, smoking, alcohol, BMI, physical activity, sleep duration. Model 3: Model 2 + diabetes, hypertension, hyperlipidemia. HR indicates hazard ratio; CI, confidence interval; CHIP, clonal hematopoiesis of indeterminate potential; VAF, variant allele frequency.*

**eTable 3. Sensitivity Analyses**

| **Analysis** | **Outcome** | **Exposure** | **HR (95% CI)** | **P-value** |
| --- | --- | --- | --- | --- |
| **2-year landmark** | Cataract | Any CHIP | 1.07 (1.02-1.13) | .01 |
|  | Glaucoma | Any CHIP | 1.10 (1.00-1.20) | .04 |
|  | AMD | Any CHIP | 1.13 (1.04-1.23) | .003 |
|  | DR | Any CHIP | 1.40 (1.20-1.65) | < .001 |
| **White only** | Cataract | Any CHIP | 1.10 (1.04-1.16) | .001 |
|  | Glaucoma | Any CHIP | 1.09 (1.00-1.20) | .06 |
|  | AMD | Any CHIP | 1.12 (1.03-1.22) | .009 |
|  | DR | Any CHIP | 1.39 (1.17-1.66) | < .001 |

*2-year landmark analysis excludes events occurring within the first 2 years of follow-up. White ethnicity only analysis restricts to participants of White European ancestry. All models use Model 3 adjustments.*

**eTable 4. Gene-Specific Association of CHIP With Incident Eye Diseases**

| **Outcome** | **Gene** | **Events** | **Model 1 HR (95% CI)** | **Model 2 HR (95% CI)** | **Model 3 HR (95% CI)** |
| --- | --- | --- | --- | --- | --- |
| **Cataract** | DNMT3A | 31,165 | 1.06 (0.99-1.14) (P = .075) | 1.06 (0.99-1.13) (P = .079) | 1.06 (0.99-1.14) (P = .073) |
|  | TET2 |  | 1.10 (0.96-1.26) (P = .177) | 1.10 (0.96-1.26) (P = .185) | 1.10 (0.96-1.27) (P = .160) |
|  | ASXL1 |  | 1.13 (0.97-1.31) (P = .106) | 1.09 (0.94-1.26) (P = .269) | 1.09 (0.94-1.26) (P = .276) |
|  | Other |  | 1.15 (1.01-1.32) (P = .041) | 1.13 (0.99-1.30) (P = .077) | 1.12 (0.98-1.29) (P = .094) |
| **Glaucoma** | DNMT3A | 12,393 | 1.09 (0.98-1.22) (P = .104) | 1.09 (0.98-1.22) (P = .108) | 1.09 (0.98-1.22) (P = .107) |
|  | TET2 |  | 1.15 (0.92-1.44) (P = .219) | 1.15 (0.92-1.43) (P = .228) | 1.15 (0.92-1.43) (P = .225) |
|  | ASXL1 |  | 0.97 (0.75-1.26) (P = .847) | 0.97 (0.75-1.26) (P = .834) | 0.97 (0.75-1.26) (P = .835) |
|  | Other |  | 1.02 (0.80-1.28) (P = .896) | 1.01 (0.80-1.28) (P = .933) | 1.00 (0.79-1.27) (P = .970) |
| **AMD** | DNMT3A | 13,644 | 1.10 (0.99-1.21) (P = .072) | 1.10 (0.99-1.21) (P = .071) | 1.10 (0.99-1.21) (P = .066) |
|  | TET2 |  | 1.06 (0.86-1.31) (P = .601) | 1.05 (0.85-1.30) (P = .631) | 1.06 (0.86-1.32) (P = .577) |
|  | ASXL1 |  | 1.19 (0.95-1.48) (P = .134) | 1.15 (0.92-1.44) (P = .214) | 1.15 (0.92-1.44) (P = .213) |
|  | Other |  | 1.27 (1.04-1.54) (P = .019) | 1.25 (1.02-1.52) (P = .028) | 1.23 (1.01-1.50) (P = .038) |
| **DR** | DNMT3A | 3,292 | 1.25 (1.01-1.53) (P = .036) | 1.29 (1.05-1.58) (P = .017) | 1.36 (1.10-1.66) (P = .004) |
|  | TET2 |  | 1.26 (0.83-1.91) (P = .284) | 1.25 (0.82-1.90) (P = .298) | 1.40 (0.92-2.12) (P = .119) |
|  | ASXL1 |  | 1.38 (0.90-2.09) (P = .136) | 1.24 (0.81-1.88) (P = .320) | 1.24 (0.82-1.89) (P = .308) |
|  | Other |  | 1.90 (1.36-2.67) (P < .001) | 1.81 (1.29-2.54) (P < .001) | 1.76 (1.27-2.45) (P < .001) |

*Model adjustments are the same as in eTable 2.*

**eTable 5. Subgroup Analyses of CHIP and Incident Eye Diseases**

| **Outcome / Subgroup** | **Stratum** | **HR (95% CI)** | **P-value** | **P-interaction** |
| --- | --- | --- | --- | --- |
| **Cataract** |  |  |  |  |
| Age, y | Age < 60 | 1.48 (1.30-1.69) | < .001 | < .001 |
|  | Age >= 60 | 1.13 (1.07-1.19) | < .001 |  |
| Sex | Female | 1.07 (1.00-1.14) | .07 | .73 |
|  | Male | 1.11 (1.02-1.20) | .02 |  |
| BMI, kg/m² | BMI < 30 | 1.09 (1.03-1.16) | .004 | .31 |
|  | BMI >= 30 | 1.05 (0.95-1.16) | .38 |  |
| Diabetes | No Diabetes | 1.07 (1.01-1.13) | .02 | .51 |
|  | Diabetes | 1.20 (1.02-1.41) | .03 |  |
| Hypertension | No Hypertension | 1.07 (0.98-1.16) | .15 | .94 |
|  | Hypertension | 1.09 (1.02-1.16) | .009 |  |
| **Glaucoma** |  |  |  |  |
| Age, y | Age < 60 | 1.35 (1.12-1.62) | .001 | .04 |
|  | Age >= 60 | 1.09 (0.99-1.21) | .07 |  |
| Sex | Female | 1.13 (1.00-1.27) | .04 | .22 |
|  | Male | 1.02 (0.90-1.16) | .75 |  |
| BMI, kg/m² | BMI < 30 | 1.12 (1.01-1.23) | .02 | .05 |
|  | BMI >= 30 | 0.96 (0.80-1.15) | .63 |  |
| Diabetes | No Diabetes | 1.06 (0.97-1.17) | .18 | .74 |
|  | Diabetes | 1.21 (0.91-1.60) | .19 |  |
| Hypertension | No Hypertension | 1.22 (1.06-1.40) | .005 | .01 |
|  | Hypertension | 1.00 (0.89-1.12) | .99 |  |
| **AMD** |  |  |  |  |
| Age, y | Age < 60 | 1.36 (1.12-1.66) | .002 | .15 |
|  | Age >= 60 | 1.18 (1.08-1.28) | < .001 |  |
| Sex | Female | 1.09 (0.98-1.21) | .10 | .64 |
|  | Male | 1.17 (1.03-1.31) | .01 |  |
| BMI, kg/m² | BMI < 30 | 1.12 (1.02-1.23) | .01 | .71 |
|  | BMI >= 30 | 1.12 (0.96-1.30) | .15 |  |
| Diabetes | No Diabetes | 1.10 (1.01-1.20) | .03 | .73 |
|  | Diabetes | 1.28 (1.03-1.59) | .03 |  |
| Hypertension | No Hypertension | 1.03 (0.90-1.19) | .63 | .20 |
|  | Hypertension | 1.17 (1.07-1.29) | .001 |  |
| **DR** |  |  |  |  |
| Age, y | Age < 60 | 1.36 (0.98-1.88) | .07 | .39 |
|  | Age >= 60 | 1.44 (1.21-1.72) | < .001 |  |
| Sex | Female | 1.55 (1.22-1.97) | < .001 | .10 |
|  | Male | 1.32 (1.08-1.61) | .007 |  |
| BMI, kg/m² | BMI < 30 | 1.22 (0.97-1.55) | .09 | .20 |
|  | BMI >= 30 | 1.56 (1.27-1.91) | < .001 |  |
| Diabetes | No Diabetes | 0.97 (0.64-1.47) | .87 | .19 |
|  | Diabetes | 1.48 (1.25-1.75) | < .001 |  |
| Hypertension | No Hypertension | 1.56 (1.15-2.13) | .005 | .06 |
|  | Hypertension | 1.35 (1.13-1.62) | < .001 |  |

*All models are adjusted for Model 3 covariates (excluding the stratification variable). P-interaction was calculated using likelihood ratio test comparing models with and without the interaction term. HR indicates hazard ratio; CI, confidence interval; BMI, body mass index.*

**eTable 6. Association of CHIP With Ocular Structural Measures**

| **Outcome** | **Exposure** | **N** | **Model 1 β (95% CI)** | **Model 2 β (95% CI)** | **Model 3 β (95% CI)** |
| --- | --- | --- | --- | --- | --- |
| **IOP (mmHg)** | Any CHIP | 102,667 | -0.066 (-0.191, 0.060) (P = .305) | -0.081 (-0.207, 0.044) (P = .203) | -0.094 (-0.218, 0.031) (P = .140) |
|  | Small CHIP |  | -0.071 (-0.283, 0.140) (P = .507) | -0.083 (-0.293, 0.128) (P = .441) | -0.083 (-0.292, 0.125) (P = .434) |
|  | Large CHIP |  | -0.063 (-0.217, 0.091) (P = .425) | -0.081 (-0.234, 0.073) (P = .304) | -0.099 (-0.251, 0.054) (P = .204) |
| **RNFL Thickness (μm)** | Any CHIP | 61,193 | -0.030 (-0.359, 0.299) (P = .857) | -0.014 (-0.342, 0.315) (P = .935) | -0.011 (-0.339, 0.318) (P = .950) |
|  | Small CHIP |  | -0.282 (-0.830, 0.265) (P = .312) | -0.276 (-0.823, 0.271) (P = .323) | -0.273 (-0.820, 0.273) (P = .327) |
|  | Large CHIP |  | 0.106 (-0.299, 0.512) (P = .607) | 0.128 (-0.277, 0.533) (P = .535) | 0.132 (-0.273, 0.537) (P = .523) |
| **Macular Thickness (μm)** | Any CHIP | 61,193 | -0.192 (-1.271, 0.886) (P = .727) | -0.126 (-1.202, 0.951) (P = .819) | -0.106 (-1.182, 0.970) (P = .847) |
|  | Small CHIP |  | -0.820 (-2.616, 0.977) (P = .371) | -0.785 (-2.578, 1.007) (P = .391) | -0.773 (-2.565, 1.019) (P = .398) |
|  | Large CHIP |  | 0.147 (-1.183, 1.478) (P = .828) | 0.232 (-1.096, 1.559) (P = .732) | 0.255 (-1.072, 1.582) (P = .706) |

*IOP indicates intraocular pressure; RNFL, retinal nerve fiber layer. β coefficients represent the difference in outcome measure associated with CHIP status. Model adjustments are the same as in eTable 2.*

**eTable 7. Gene-Specific Association of CHIP With Ocular Structural Measures**

| **Outcome** | **Gene** | **Model 1 β (95% CI)** | **Model 2 β (95% CI)** | **Model 3 β (95% CI)** |
| --- | --- | --- | --- | --- |
| **IOP (mmHg)** | DNMT3A | -0.009 (-0.166, 0.149) (P = .913) | -0.015 (-0.172, 0.142) (P = .848) | -0.027 (-0.183, 0.129) (P = .733) |
|  | TET2 | -0.365 (-0.711, -0.018) (P = .039) | -0.376 (-0.721, -0.031) (P = .033) | -0.388 (-0.730, -0.045) (P = .027) |
|  | ASXL1 | 0.150 (-0.233, 0.532) (P = .443) | 0.078 (-0.303, 0.459) (P = .689) | 0.092 (-0.285, 0.470) (P = .632) |
|  | Other | -0.191 (-0.518, 0.135) (P = .251) | -0.205 (-0.531, 0.120) (P = .216) | -0.237 (-0.560, 0.085) (P = .149) |
| **RNFL Thickness (μm)** | DNMT3A | -0.005 (-0.416, 0.406) (P = .981) | -0.019 (-0.429, 0.391) (P = .928) | -0.018 (-0.428, 0.392) (P = .933) |
|  | TET2 | 0.209 (-0.701, 1.120) (P = .652) | 0.220 (-0.689, 1.129) (P = .635) | 0.211 (-0.698, 1.119) (P = .649) |
|  | ASXL1 | -0.495 (-1.526, 0.536) (P = .346) | -0.407 (-1.436, 0.622) (P = .438) | -0.382 (-1.411, 0.648) (P = .467) |
|  | Other | -0.026 (-0.878, 0.825) (P = .952) | 0.075 (-0.775, 0.925) (P = .863) | 0.081 (-0.769, 0.931) (P = .852) |
| **Macular Thickness (μm)** | DNMT3A | 0.239 (-1.108, 1.587) (P = .728) | 0.208 (-1.136, 1.553) (P = .762) | 0.216 (-1.128, 1.559) (P = .753) |
|  | TET2 | -0.160 (-3.144, 2.825) (P = .917) | -0.051 (-3.030, 2.928) (P = .973) | -0.078 (-3.055, 2.900) (P = .959) |
|  | ASXL1 | -1.596 (-4.976, 1.784) (P = .355) | -1.362 (-4.736, 2.012) (P = .429) | -1.241 (-4.614, 2.131) (P = .471) |
|  | Other | -1.089 (-3.881, 1.703) (P = .445) | -0.763 (-3.549, 2.024) (P = .592) | -0.722 (-3.507, 2.064) (P = .611) |

*Model adjustments are the same as in eTable 2.*

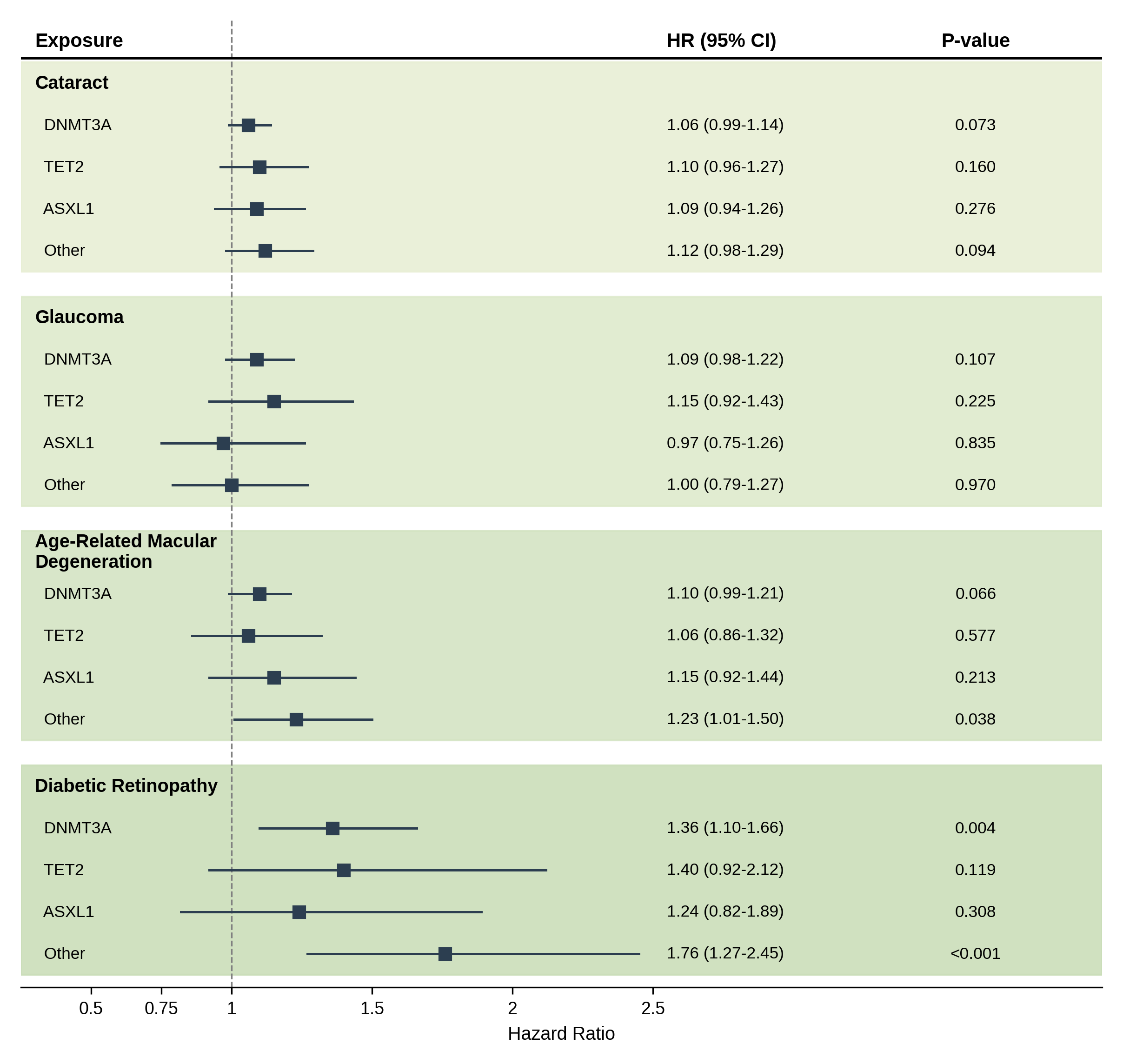

**eFigure 1. Gene-Specific Associations of CHIP with Incident Eye Diseases**

*Forest plot showing hazard ratios (HRs) and 95% CIs for the associations of CHIP, stratified by driver gene (DNMT3A, TET2, ASXL1, and Other), with incident cataract, glaucoma, age-related macular degeneration (AMD), and diabetic retinopathy (DR). Results are from the fully adjusted Cox proportional hazards model (Model 3).*

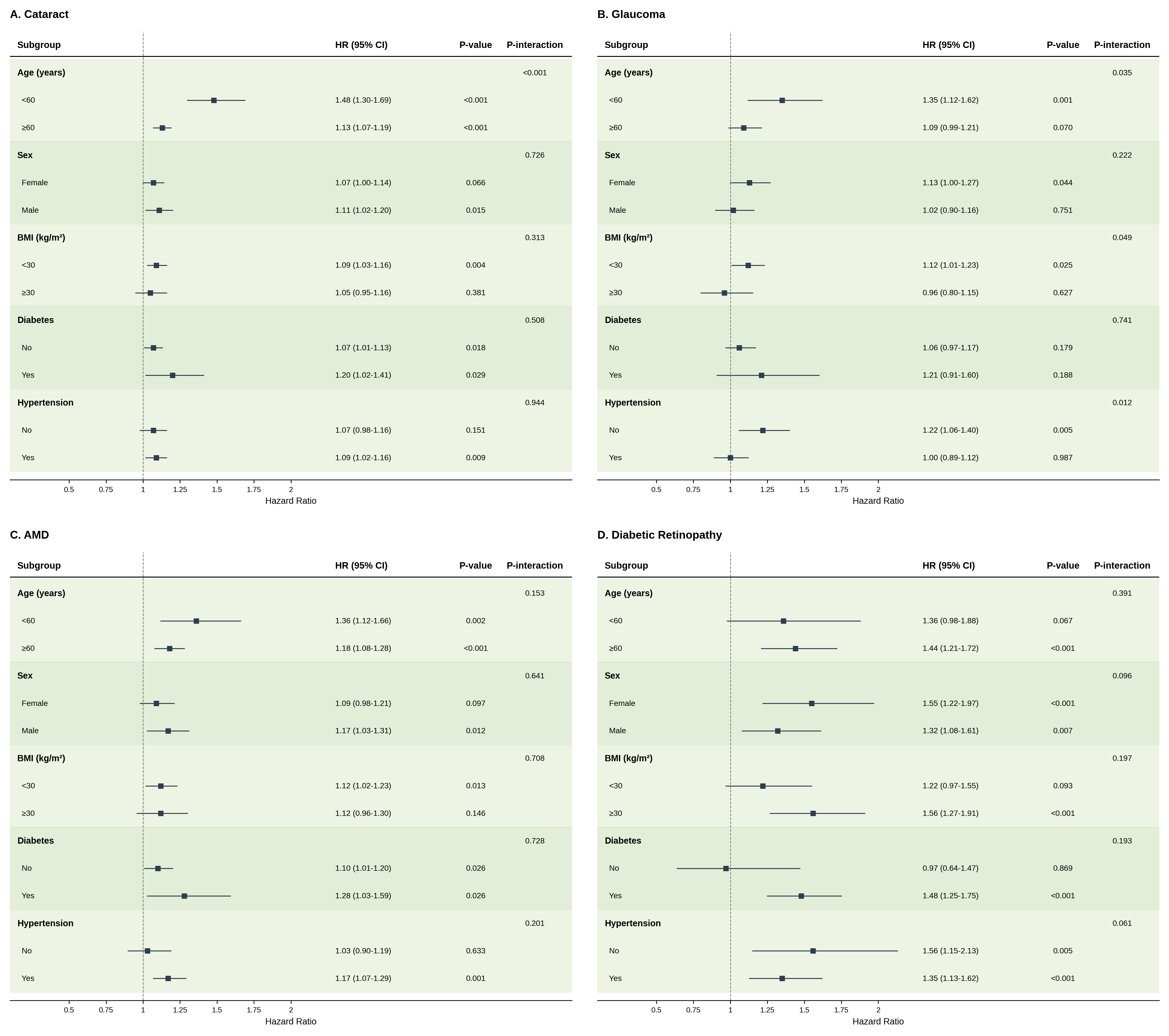

**eFigure 2. Subgroup Analyses of the Association Between CHIP and Incident Eye Diseases**

*Forest plot showing hazard ratios (HRs) and 95% CIs for the association between any CHIP and incident eye diseases across pre-specified subgroups: age (<60 vs ≥60 years), sex (female vs male), body mass index (<30 vs ≥30 kg/m²), prevalent diabetes (no vs yes), and prevalent hypertension (no vs yes). P-values for interaction were derived from multiplicative interaction terms in the fully adjusted Cox model.*
